# How did labelling provision on menus for online food delivery change following England’s calorie labelling regulations?

**DOI:** 10.1101/2025.09.26.25336607

**Authors:** Alexandra Kalbus, Oana-Adelina Tanasache, Cherry Law, Jean Adams, Penny Breeze, Kerry Ann Brown, Steven Cummins, Dalya Marks, Stephen O’Neill, Richard Smith, Laura Cornelsen

## Abstract

This study assessed the provision of calorie labelling in the digital out-of-home food sector following the implementation of mandatory calorie labelling for large businesses in England in April 2022. Using online menu data from two major food delivery services between June 2022 and October 2023, the number of restaurants available for delivery and the share of restaurants displaying calories was determined for every neighbourhood (Lower layer Super Output Area/Data Zone) in Great Britain. Among restaurants that display calories, we determined the share of labelled menu items and these items’ calorie content. We assessed changes by area deprivation and restaurant type. Online food delivery was available in ∼89% of Great Britain, with more restaurants available in more deprived neighbourhoods. The share of restaurants that display calories was overall low and decreasing over time (median 14% in June 2022 to 12% in October 2023), and was lowest in most deprived (9%) compared with 14% in the least deprived neighbourhoods in October 2023. Among restaurants displaying calories, the share of labelled items was high (79%–76%), while calorie content decreased slightly (by 14 kcal/food item and 5 kcal/drink item). Changes by deprivation were limited and heterogenous by restaurant type. The observed reduction in median calorie content may suggest positive, structural change and warrants further investigation. However, due to poor coverage calorie labelling is limited, and dietary health inequalities may widen if it continues to be more likely in more affluent neighbourhoods.

## Introduction

As part of policy efforts to address high levels of poor dietary health, mandatory calorie labelling was introduced in England, UK, in 2022 (UK Government, 2021). The calorie labelling regulations require large businesses (those with 250 or more employees) in the out-of-home (OOH) food sector such as restaurants, cafes, and takeaways, to show calories (in kcal) on their menus. Calories must be displayed at the point of choice, which includes physical and online menus, for food and non-alcoholic drink items prepared for immediate consumption. Some exceptions apply, such as loose fresh produce or items that are on the menu for less than 30 consecutive days (UK Government, 2021).

Calorie labelling is thought to reduce calories consumed in two ways (Zlatevska et al., 2018): First, consumers may utilise the nutritional information provided to purchase and consume fewer calories. Second, calorie disclosure may encourage businesses to lower the overall calorie content of the menu through changing the foods and drinks they offer.

Mandatory calorie disclosure has been implemented in several countries worldwide, including the US, Canada, and Australia (Rincón-Gallardo Patiño et al., 2020). To date, evidence of the efficacy of calorie labelling is mixed. Regarding the first mechanism of consumer behaviour change, an overall reduction of 11 kcal per meal selected has been reported by a recent systematic review and meta-analysis (Clarke et al., 2025). However, this estimate was predominantly informed by experimental studies set in the US context. In England, no change in consumer behaviour following the introduction of calorie labelling regulations has been observed (Kalbus et al., 2025; Polden et al., 2024b).

The evidence base with respect to menu change as a second mechanism, albeit inconsistent, points to small decreases in calories following mandatory disclosure. A meta-analysis of US studies suggests that calorie labelling was associated with a reduction of 15 kcal per menu item on average (Zlatevska et al., 2018). In contrast, research from Canada (Scourboutakos et al., 2019) and Australia (Wellard-Cole et al., 2018) did not find changes in calories on menus following mandatory calorie labelling. In England, there have been two studies to date on changes in calories following the calorie labelling regulations. Luick et al. examined menu changes in workplace canteens and observed decreases in calorie content with each changing menu, e.g. -4.2 kcal per item in July 2022 compared to the pre-policy period (2024). Analysing menu items from 78 large restaurant chains, Essman et al. reported a mean reduction of 9 kcal per menu item between pre- and post-policy implementation (2024).

However, an important gap in the evidence to date is that it does not cover the digital OOH food sector. Conceptualised as third-party aggregator platforms such as Uber Eats, which facilitate delivery of food and drink items from food businesses to customers, the digital OOH food sector has expanded considerably in recent years (Kalbus et al., 2023; Vanderlee and Sacks, 2023). More than half of the UK population (55%) report to have ever ordered from an online food delivery platform (Moore et al., 2025). Of those who have ordered through these platforms, most report to have done so 2–3 times a month or less often (74%), while 21% indicated ordering about once a week or more often (Moore et al., 2025). Previous research suggests that the use of online takeaways is associated with lower socio-economic status and greater likelihood of living with obesity (Cornelsen et al., 2025; Cummins et al., 2024). Despite its public health relevance (Greenthal et al., 2023; Vanderlee et al., 2023a), no study to date has investigated calorie labelling in the context of online food delivery in Great Britain.

We address this gap by assessing calorie labelling among the digital OOH food sector after the implementation of mandatory calorie labelling. We focus on Great Britain despite the calorie labelling regulations only applying to England as many large businesses operate across borders to examine calorie labelling provision overall and by region. Specifically, the study seeks to understand any changes during the first year post-policy implementation in the prevalence of restaurants displaying calories on their online menus (analysis 1); and for the restaurants that display calories, any changes in the share of menu items with calorie information (analysis 2); and in the calorie content of menu items (analysis 3). To understand implications on dietary inequalities, we stratified analyses by area deprivation and restaurant type. Our study complements existing evidence in understanding the reach of the policy and potential inequalities in exposure to labelling, thus providing stakeholders with crucial information on real-world policy effects.

## Methods

### Design

We undertook a descriptive, longitudinal analysis of online menu data for Great Britain between June 2022 and October 2023.

### Data

Online menu data were automatically collected (web-scraped) from the websites of Deliveroo and Uber Eats, two major UK online food delivery platforms. Their market share in 2023 was 25% for Deliveroo and 20% for Uber Eats (SLERP, 2024). Bespoke web-scraping tools (Greener, 2023, 2022) collected all public-facing information on the restaurant page, including name, location (coordinates), tags, menu item names and descriptions, price, and calorie content where this information was available (labelled items). Data were collected using as input the geographic centroid for each Lower layer Super Output Area (LSOA) in England (n=32,844) and Wales (n=1,909), and for each Data Zone in Scotland (n=6,976) using 2011 boundaries. LSOAs and Data Zones, hereafter referred to as neighbourhoods, are geographies used in the census, and typically comprise between 1,000 and 3,000 (LSOAs) and between 500 and 1,000 (Data Zones) usual residents. Data were collected at five points in time: June 2022, October 2022, February 2023, June 2023, and October 2023. For ease of presentation, the latter refers to Uber Eats data from August 2023 and Deliveroo data from October 2023.

#### Data preparation

Data were prepared according to the analyses shown in Table 1.

**Table 1.**
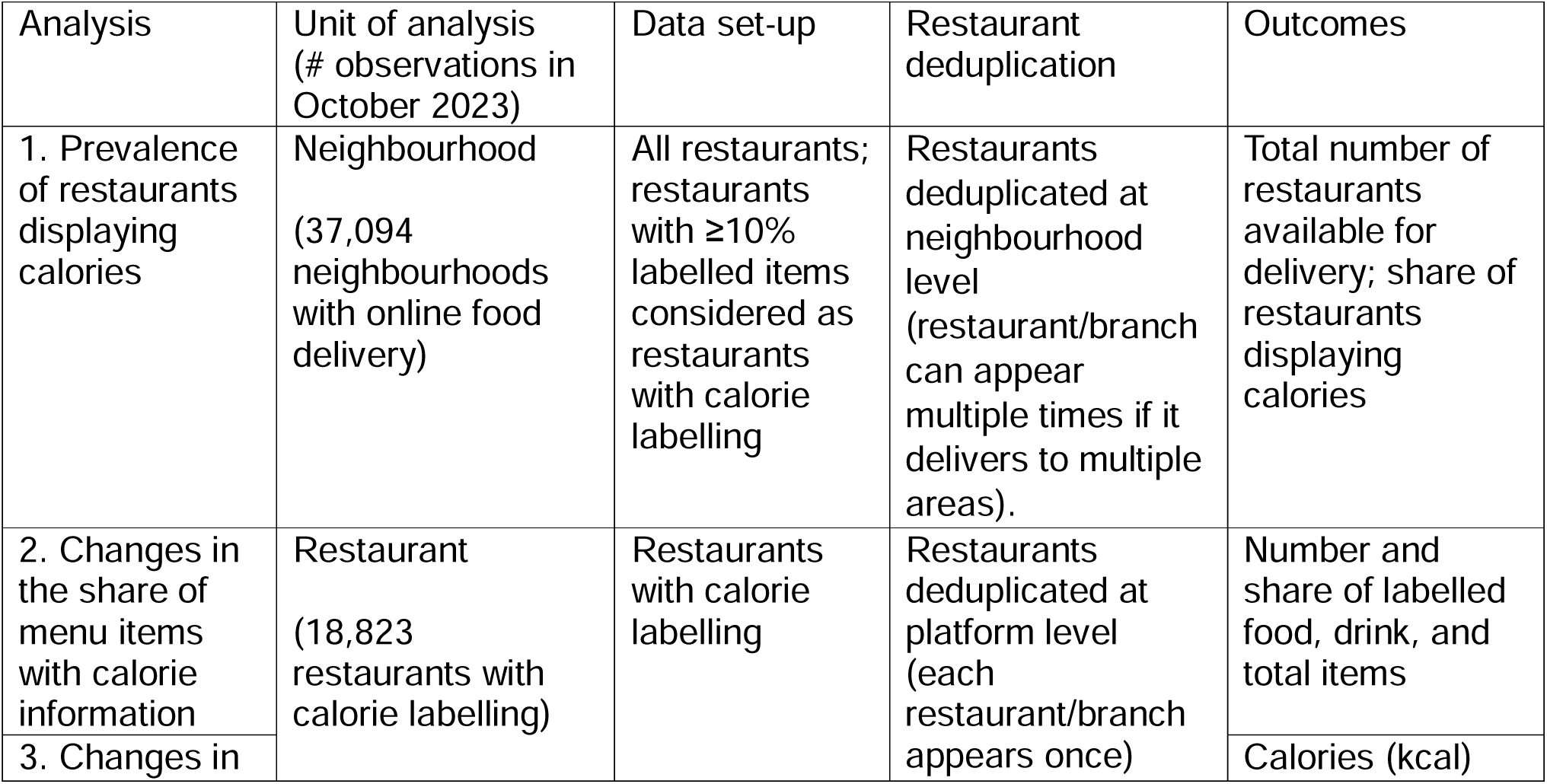
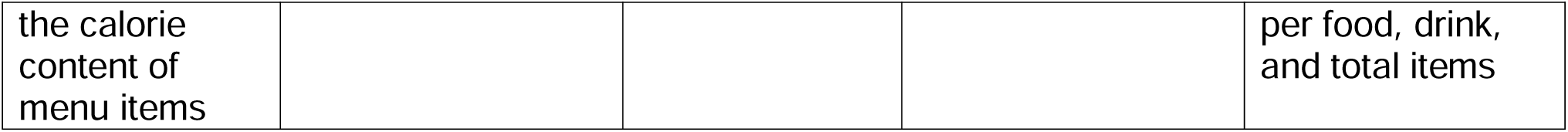
Data preparation according to the analyses.

| Table 1. Data preparation according to the analyses |  |  |  |  |
| --- | --- | --- | --- | --- |
| Analysis | Unit of analysis<br>(# observations in<br>October 2023) | Data set-up | Restaurant<br>deduplication | Outcomes |
| 1. Prevalence<br>of restaurants<br>displaying<br>calories | Neighbourhood<br>(37,094<br>neighbourhoods<br>with online food<br>delivery) | All restaurants;<br>restaurants<br>with ≥10%<br>labelled items<br>considered as<br>restaurants<br>with calorie<br>labelling | Restaurants<br>deduplicated at<br>neighbourhood<br>level<br>(restaurant/branch<br>can appear<br>multiple times if it<br>delivers to multiple<br>areas). | Total number of<br>restaurants<br>available for<br>delivery; share of<br>restaurants<br>displaying<br>calories |
| 2. Changes in<br>the share of<br>menu items<br>with calorie<br>information | Restaurant<br>(18,823<br>restaurants with<br>calorie labelling) | Restaurants<br>with calorie<br>labelling | Restaurants<br>deduplicated at<br>platform level<br>(each<br>restaurant/branch<br>appears once) | Number and<br>share of labelled<br>food, drink, and<br>total items |
| 3. Changes in |  |  |  | Calories (kcal) |
| the calorie content of menu items |  |  |  | per food, drink, and total items |

We removed grocery, retail and non-food outlets, leaving restaurants, takeaways, cafes, pubs, as well as delivery only (i.e. dark kitchens) which may or may not belong to high street chains, collectively referred to as ‘restaurants’. We deduplicated restaurants available through both platforms. A detailed description of the deduplication process is provided in Appendix A. In brief, record pairs, i.e. pairs of one restaurant from each platform, were removed if they were closer than 300 m apart and their names were similar based on the Optimal String Alignment distance.

We prepared the merged item-level dataset by removing non-food and items not intended for immediate consumption on their own, e.g. dips, condiments, sauces and toppings. We distinguished between food and drink items and extracted calorie (kcal) information where available. We defined ‘restaurants displaying calories’ as restaurants with calorie information for at least 10% of menu items, which data exploration revealed to be a cut-off between large food businesses and smaller restaurants displaying calories inconsistently, e.g. only for a few pre-packaged food or drink items. We defined sharing items as those declared as for sharing (e.g. ‘family feast’) and/or exceeding 2,500 kcal, corresponding to the UK daily energy intake guideline for an average man (Public Health England, 2016), and transformed their calorie content to reflect one serving as indicated in the data. Sharing items for which the serving size was unknown were excluded.

#### Additional data

Neighbourhood-level data were supplemented with information on area deprivation, population density, and urban status. We determined each neighbourhood’s relative deprivation through the Townsend Index calculated from the 2021 (England and Wales) and 2022 (Scotland) census (Office for National Statistics, 2022a; Scotland’s Census, 2024) using the look-up tool between 2021 and 2011 boundary data (Office for National Statistics, 2022b), and derived quintiles of deprivation (Yousaf and Bonsall, 2017). Population density was calculated as the number of usual residents per km^2^ using population estimates and area boundaries from the most recent census (Office for National Statistics, 2023a, 2022a; Scotland’s Census, 2024; Scottish Government, 2014). Urban status was available for England and Wales from 2011 (Office for National Statistics, 2018), and for Scotland from 2020 (Scottish Health and Social Care Open Data, 2020).

Data cleaning was carried out in Python and R version 4.5.0, while the analysis was performed in R.

### Analysis

Descriptive analysis includes summary statistics and boxplots. Due to skewed distributions, medians and interquartile ranges (IQR) are presented throughout.

#### Analysis 1

We computed the median and IQR of the total number and share of restaurants displaying calories over time and by area deprivation. Using the *leaflet* package

(Cheng et al., 2024) and neighbourhood boundaries (Office for National Statistics, 2023b; Scottish Government, 2024), we created interactive maps of the prevalence of restaurants with calorie labelling across Great Britain and Greater London. Further analysis explored whether observed relationships between area deprivation and labelling provision differed by region and urban status. Additionally, we used a linear regression model following the form: Provision of calorie labelling per neighbourhood = area deprivation quintile + region + area deprivation quintile x region + population density + urban status. We also predicted the total number of restaurants available for delivery per neighbourhood using the same model specification as described above. Using the *marginaleffects* package (Arel-Bundock et al., 2024), we computed adjusted average predictions of the total number of restaurants available for delivery and calorie labelling provision per deprivation quintile. Models used data from October 2023 as the most recent data available.

#### Analyses 2 and 3

We calculated the median and IQR of the number and share of labelled food, drink and all items, as well as median and IQR of kcal per menu item, weighted by the number of neighbourhoods each restaurant delivers to. We explored differences by area-level deprivation of the neighbourhood which each restaurant delivers to. In doing so, we considered each neighbourhood-restaurant combination, effectively weighting the data by the number of neighbourhoods each restaurant delivered to, and calculated the median share of labelled items and median kcal per item for each area deprivation quintile. To understand whether changes in calorie content were due to reformulation of existing dishes or changes to menus, we also stratified by whether an item was continuously on the menu (i.e. present in June 2022 and October 2023) or not.

#### Secondary analysis

Secondary analysis explored each research question by restaurant type as previous research indicated variation by cuisine type (Essman et al., 2024). The latter was based on the 11 most popular tags self-defined by restaurants (∼68% of all restaurants, ∼90% of restaurants displaying calories), namely ‘Burger’, ‘British’, ‘Breakfast & Brunch’, ‘Café, ‘Family’, ‘Healthy’, ‘Italian’, ‘North American’, ‘Sandwich’, ‘Sweets & Dessert’, and ‘Vegan/Vegetarian’. Restaurant type could not be consistently determined for October 2023, when one of the platforms changed the way tags were recorded. Therefore, this analysis considered data up until June 2023.

## Results

### Analysis 1: Prevalence of restaurants displaying calories

Between June 2022 and October 2023, online food delivery services were available in the majority of neighbourhoods in Great Britain (88.4–89.9%). The number of restaurants listed on one or both included delivery service platforms increased by 16% from 98,289 in June 2022 to 114,256 in October 2023 (see Appendix B, Table B.1).

Although the median number of restaurants displaying calories per neighbourhood increased slightly from 27 (IQR 10 to 52) restaurants in June 2022 to 29 (IQR 9 to 61) restaurants in October 2023, the share of restaurants displaying calories decreased from 14.0% (IQR 8.8 to 20.9) in June 2022 to 12.1% (IQR 7.9 to 18.0) in October 2023, following a steady downward trend (see Figure 1 and Appendix B, Table B.1).

**Figure 1.**
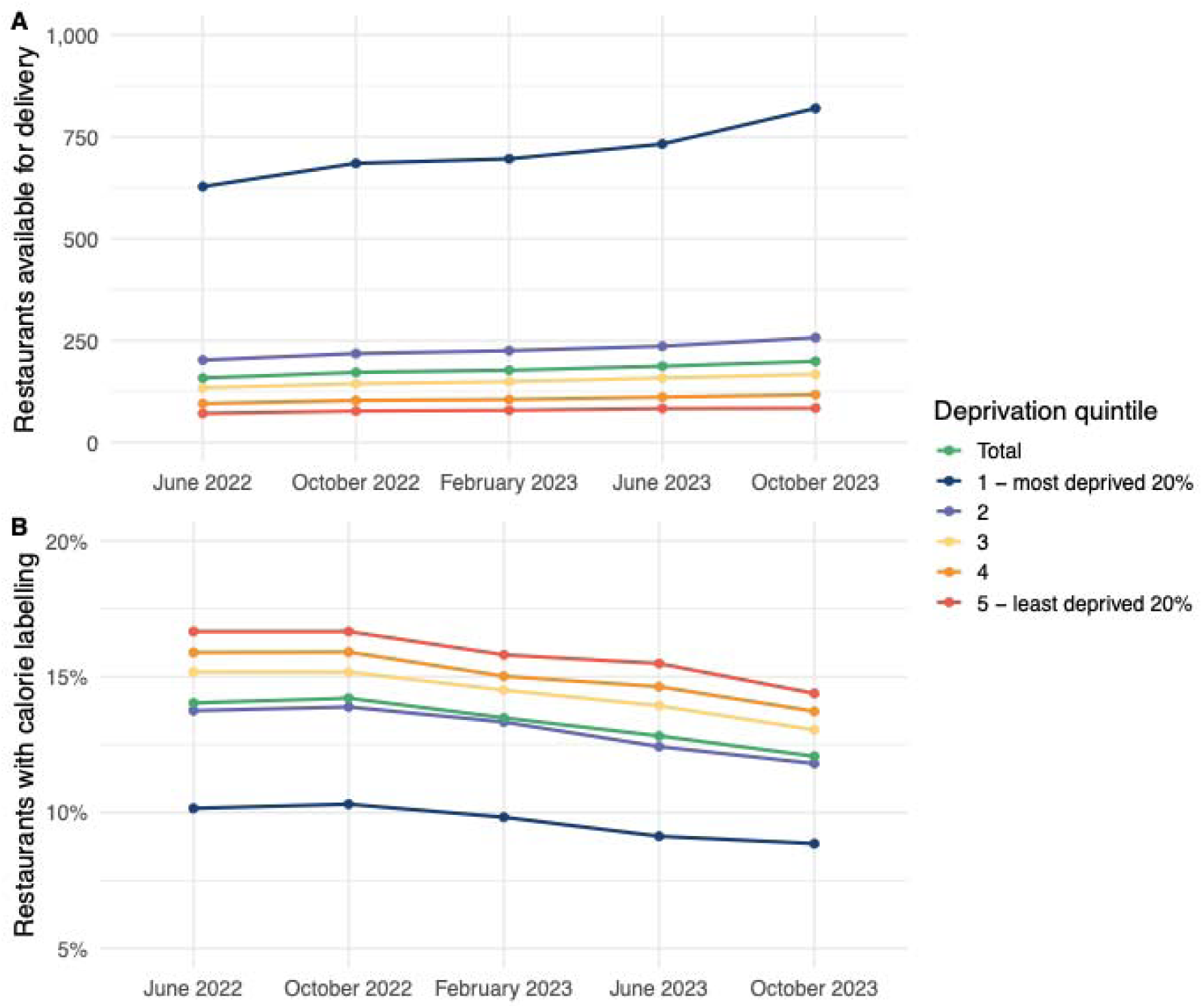
Number of restaurants available for delivery (A) and share of restaurants displaying calories (B) over time and by area deprivation. Area deprivation was approximated through the Townsend Index. Number of restaurants and % restaurants with calorie labelling are expressed as median number of all restaurants and percentage of restaurants delivering to a neighbourhood (LSOA in England & Wales/Data Zone in Scotland), respectively. The share of restaurants displaying calories varied by region, with the highest median share observed in the South West of England (17.6%, IQR 12.1 to 20.7) and the lowest in London (6.9%, IQR 6.1 to 7.7). These differences remained stable over the study period (see Appendix B, Table B.2). Despite being compulsory only in England, calorie labelling provision was comparable in England, Scotland and Wales (12.1%, 14.2%, 13.7% in October 2023, respectively). The observed inverse relationship between area deprivation and share of restaurants displaying calories differed by region, with this relationship more pronounced in English regions compared to Scotland and Wales, except in the North East and the East Midlands (see Figure 2).

**Figure 2.**
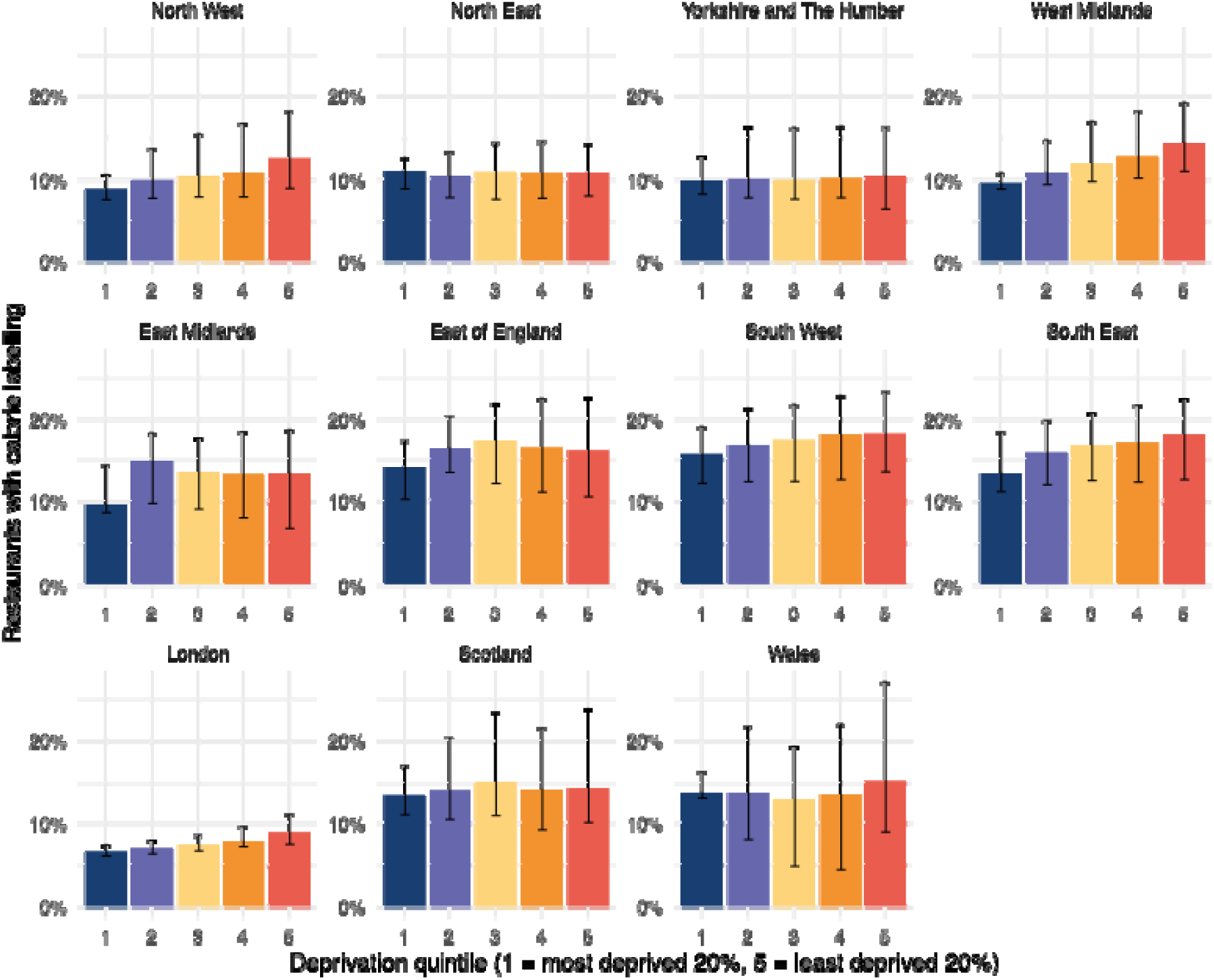
Share of restaurants displaying calories by area deprivation and region in October 2023. % restaurants displaying calories denotes the median percentage of restaurants delivering to a neighbourhood (LSOA in England and Wales/Data Zone in Scotland). Area deprivation was approximated through the Townsend Index. Error bars indicate the interquartile range.

#### Calorie labelling provision by area deprivation

While more deprived neighbourhoods had higher numbers of restaurants available for delivery overall, the share of restaurants displaying calorie labels was lower compared to less deprived neighbourhoods (see Figure 1). For example, in October 2023, the median number of restaurants delivering to a neighbourhood in the most deprived quintile was 820 (IQR 294 to 1,633), with 8.9% (IQR 6.8 to 12.5) of these displaying calories, compared with 84 (IQR 21 to 204) restaurants and 14.4% (IQR 9.3 to 20.2) in the least deprived neighbourhoods.

The relationship between deprivation and share of restaurants displaying calories was similar in urban and rural areas (see Appendix B, Figure B.1).

When adjusting for other area characteristics, the observed pattern of higher overall outlet numbers but lower share of restaurants displaying calories among more deprived neighbourhoods remained (see Appendix B, Figure B.2).

Interactive maps showing the share of restaurants displaying calories across Great Britain and London only are provided in Appendix C and D, respectively.

#### Calorie labelling provision by restaurant type

The share of restaurants displaying calories was greater among the 11 investigated restaurant types than overall, but differences between restaurant types remained stable over time (see Appendix B, Figure B.3 and Table B.3). The share of restaurants displaying calories was highest among ‘Healthy’ restaurants with a median of 45.5% (IQR 30.0 to 66.7) in June 2023, and lowest among ‘Italian’ restaurants, of which a median of 13.6% (IQR 7.0 to 27.3) displayed calories, followed closely by using ‘Burger’, ‘Café, and ‘British’ restaurants (18.2% (IQR 11.0 to 27.7), 18.2% (IQR 10.0 to 30.6)) and 18.5% (IQR 11.6 to 28.6), respectively).

### Analysis 2: Share of labelled items

The weighted median number of menu items per restaurant increased from 62 (IQR 36 to 90) in June 2022 to 67 (IQR 40 to 95) in October 2023. In contrast, the number and share of labelled items dropped. In June 2022, calorie information was provided for a median of 46 (IQR 24 to 74) menu items, decreasing slightly to 44 (IQR 23 to 80) menu items in October 2023 (see Figure 3). This corresponds to a reduction in the median share from 79.4% (IQR 63.4 to 96.7) to 76.3% (IQR 56.1 to 93.2) labelled items (see Appendix B, Table B.4).

**Figure 3.**
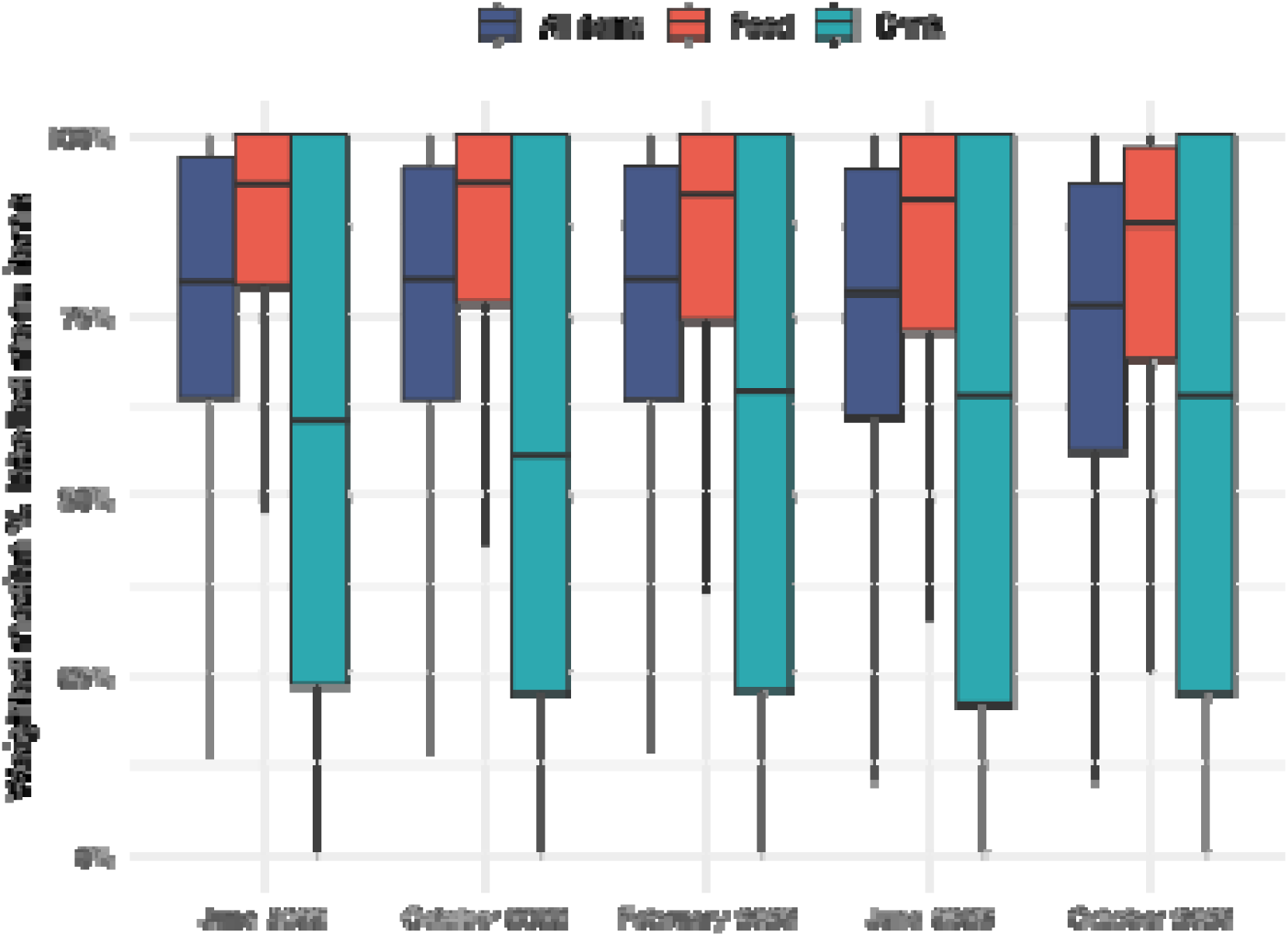
Share of labelled menu items per restaurant over time. The share was calculated as median per restaurant weighted by the number of neighbourhoods (LSOA in England and Wales/Data Zone in Scotland) the restaurants delivers to. Only restaurants displaying calories were considered. There was little variation in the share of labelled menu items across area deprivation quintiles (see Figure 4 and Appendix B, Table B.5).

**Figure 4.**
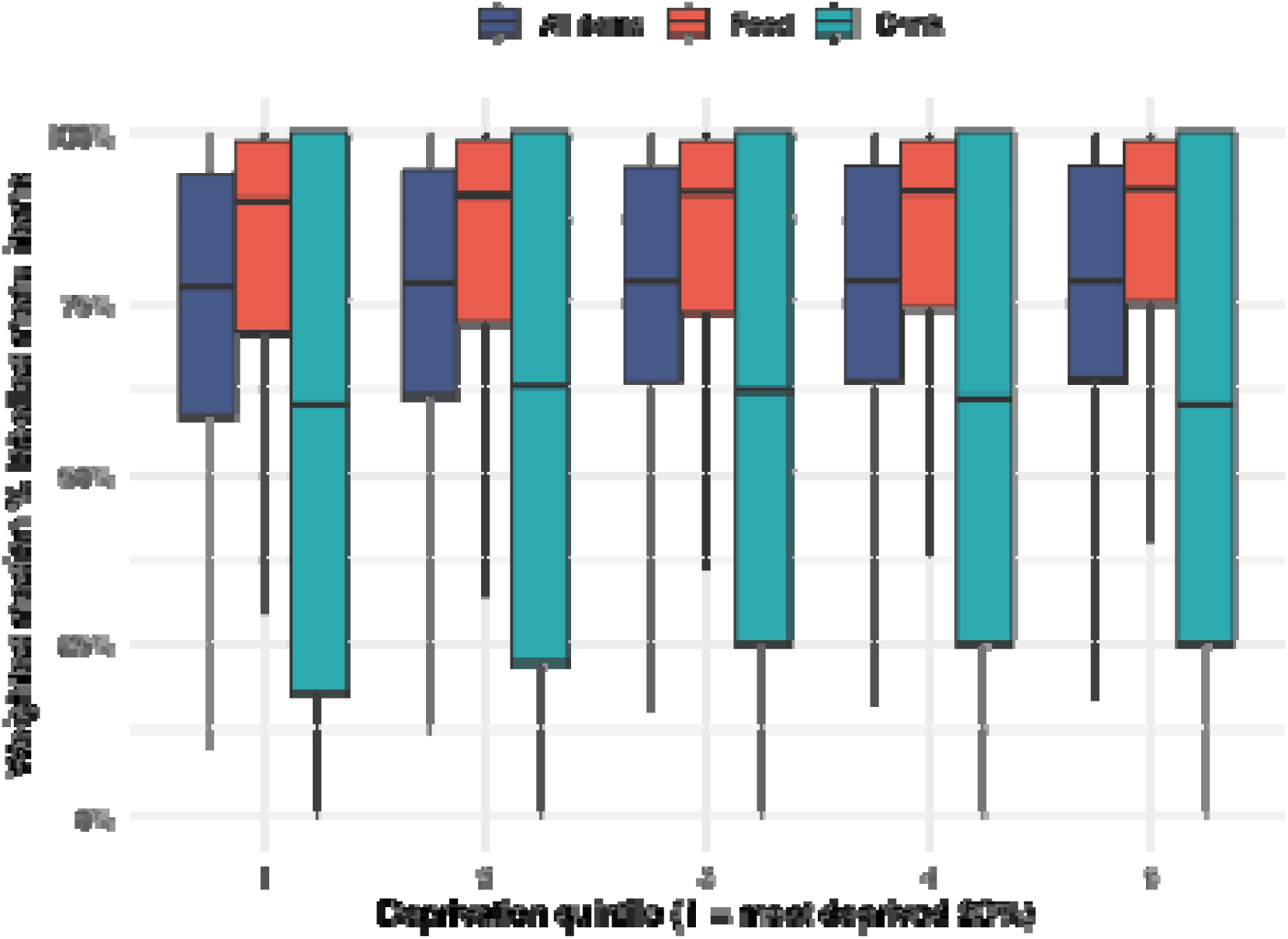
Share of labelled menu items per restaurant by area deprivation in October 2023. The share was calculated as median per restaurant weighted by the number of neighbourhoods (LSOA in England and Wales/Data Zone in Scotland) the restaurants delivers to. Only restaurants displaying calories were considered. Area deprivation was approximated through the Townsend Index.

This trend was driven by a decrease of 5.6 percentage points in the share of labelled food items from 93.1% (IQR 78.9 to 99.3) in June 2022 to 87.5% (IQR 68.6 to 98.1) in October 2023. In contrast, the share of drink items increased over time by 4.2 percentage points from 59.8% (IQR 23.2 to 98.0) in June 2022 to 63.6% (IQR 20.6 to 98.7). The absolute number of labelled food and drink items remained stable over time (see Appendix B, Table B.4).

There was some variation in the proportion of labelled menu items by restaurant type (see Appendix B, Table B.6). ‘Burger’ restaurants had the highest median share of labelled food items at 98.6% (IQR 87.9 to 99.4), while ‘Italian’ restaurants had the lowest share of 84.6% (IQR 64.7 to 92.3) in June 2023. The overall pattern of decreasing proportions of labelled food items was observed among most restaurant types.

### Analysis 3: Calorie content

The median weighted calorie content of food items decreased from 505 kcal/food item (IQR 386 to 695) in June 2022 to 491 kcal/food item (IQR 371 to 643) in October 2023, a reduction of 14 kcal (2.8%) (see Figure 5). The calorie content of drinks also decreased, from 63 kcal/drink (IQR 27 to 95) in June 2022 to 58 kcal/drink (IQR 14 to 95) in October 2023, a reduction of 5 kcal (7.5%). Narrowing IQRs over time indicate that calorie content became more homogeneous over time, with the shortening of box-plot whiskers also indicating the removal of higher-calorie dishes. This pattern was similar in menu items that were continuously on the menu and those that were not (see Appendix B, Table B.7).

**Figure 5.**
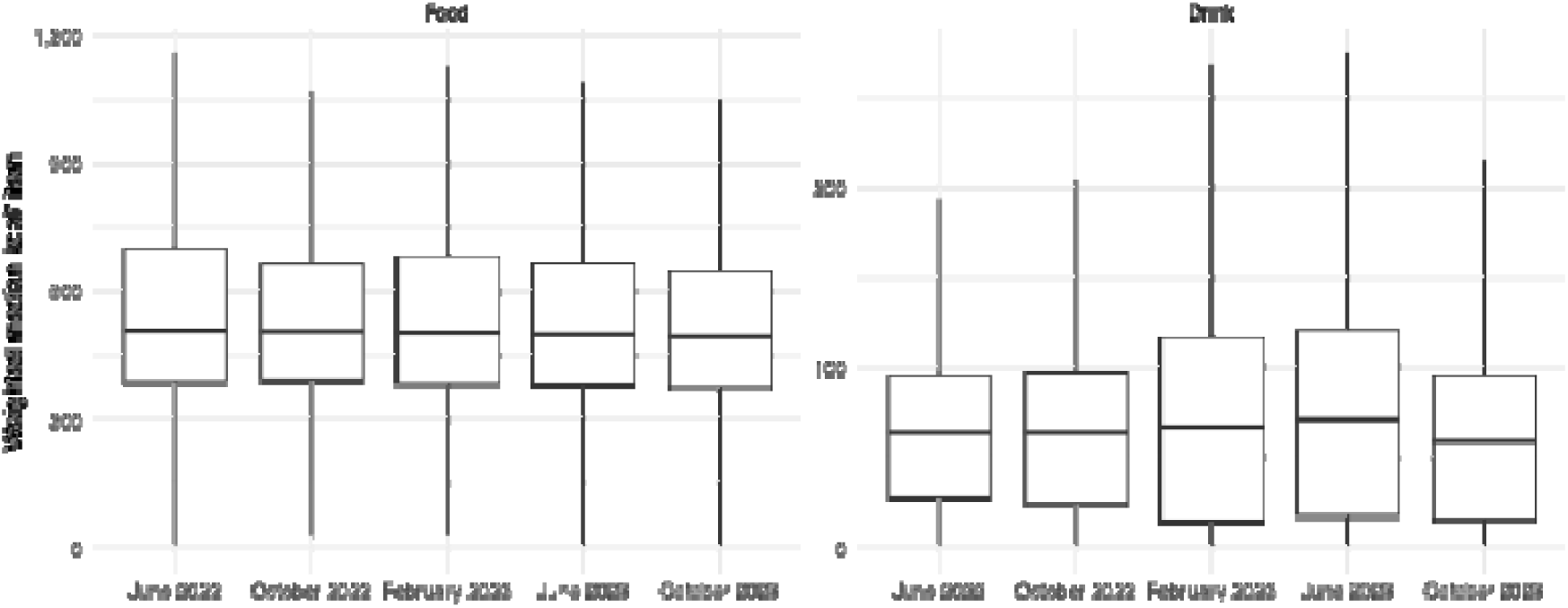
Median calorie content of food and drink items per restaurant. Medians were weighted by the number of neighbourhoods (LSOA in England and Wales/Data Zone in Scotland) the restaurant delivers to. Only restaurants displaying calories were considered.

#### Change in calorie content over time by area deprivation

Menu items’ median calorie content was similar across deprivation quintiles in June 2022 and decreased slightly in all quintiles (see Figure 6). The extent of the decrease in the calorie content of food items varied somewhat by deprivation, with a slightly higher reduction observed in the most deprived neighbourhoods (see Appendix B, Table B.8).

**Figure 6.**
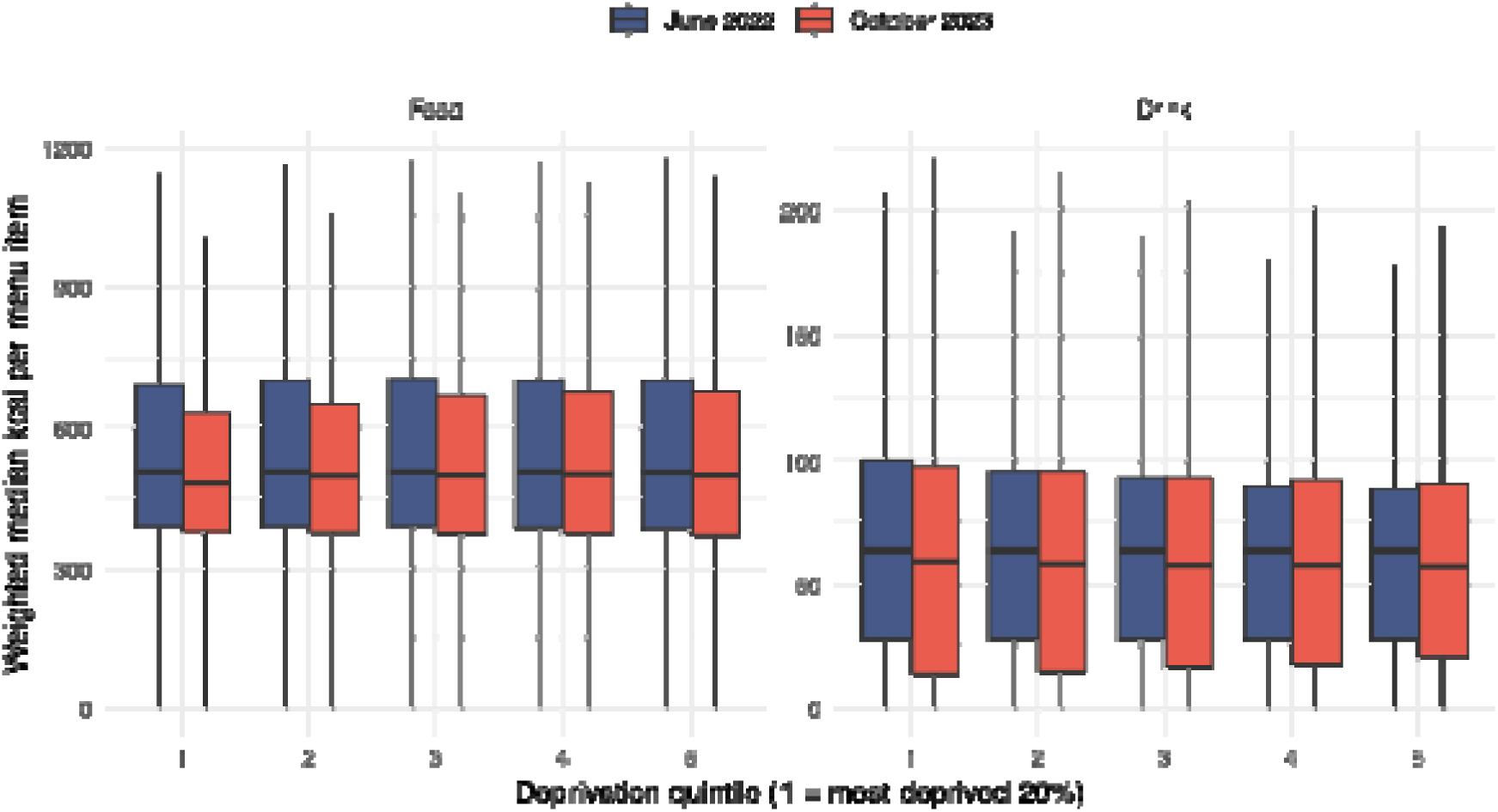
Median calorie content of food and drink items per restaurant. Medians were weighted by the number of neighbourhoods (LSOA in England and Wales/Data Zone in Scotland) the restaurant delivers to. Only restaurants displaying calories were considered. Area deprivation was approximated through the Townsend Index.

#### Change in calorie content over time by restaurant type

Also following this pattern, the calorie content of food items in most of the analysed restaurant types decreased, with reductions ranging from 8 kcal (’Healthy’ restaurants) to 87 kcal per item (‘British’ restaurants). The exception were restaurants of the type ‘North American’, ‘Breakfast and brunch’, and ‘Sandwiches’, where increases in the median calorie content were observed (+ 10 kcal/1.8%, + 4 kcal/1.1%, and + 11 kcal/3.1%, respectively). Calorie changes in drink items were less consistent by restaurant type, with the median calorie content increasing in six of the eleven types analysed by between 2 and 16 kcal per item (see Appendix B, Table B.9).

## Discussion

### Summary of findings

This study investigated changes in the provision of calorie labelling in the digital OOH food sector following the implementation of mandatory calorie labelling in England using online menu data. Between June 2022 and October 2023, the share of restaurants displaying calories was low and decreasing over time to a median of 12.1%. Calorie labelling provision was correlated with area deprivation, with a lower share of restaurants that deliver to more deprived areas providing calorie labelling. Among restaurants displaying calories, the share of labelled items was high despite a slight decrease over time. The median calorie content per food item decreased over time by 14 kcal (2.8%), while drinks decreased by 5 kcal (7.5%). There were limited differences in the share of labelled items and median calorie content by area deprivation as well as limited differences in all outcomes by restaurant type.

### Comparison with earlier research

The observed higher number of food outlets available for delivery in the most deprived 20% of neighbourhoods corresponds to the concentration of brick-and-mortar restaurants and takeaways in more deprived areas (UK Government, 2025). Findings from a recent study (Hoenink et al., 2025) linking outlets from an online food delivery platform to records of physical food outlets help explain the greater difference observed for online outlets: In more deprived areas, there is a higher proportion not only of restaurants also available online, but also of online-only restaurants such as ‘dark kitchens’ (Huang et al., 2025a), amplifying access to predominantly unhealthy food (Hoenink et al., 2025; Huang et al., 2024).

To the best of our knowledge, our study is the first to examine the provision of calorie labelling over time, space and deprivation in Great Britain. Previous international research assessed the implementation of calorie labelling among restaurants that are legally required to display calorie information (Cassano et al., 2024; Greenthal et al., 2023), but no previous work has assessed the exposure to calorie labelling overall in the digital OOH food sector.

Our findings align with emerging evidence from England which suggests decreases in calorie content following the implementation of the calorie labelling regulations (Essman et al., 2024; Luick et al., 2024). The international evidence on menu changes following calorie labelling remains mixed (Scourboutakos et al., 2019; Wellard-Cole et al., 2018; Zlatevska et al., 2018), which may be due to differences in the legislations’ scope and implementation (Rincón-Gallardo Patiño et al., 2020).

### Interpretation

The provision of calorie labelling observed in the digital OOH food sector was low and decreasing over time, suggesting limited and diminishing reach of the policy. Potential beneficial impacts of the policy on consumer behaviour may therefore be restricted to those 12% restaurants in the digital OOH food sector which display calories. The resulting low exposure to calorie labels may explain the low percentage of individuals who say that they notice calorie labels. As only 26% of those who notice calorie information (23%) further use it to reduce calories ordered online (Cornelsen et al., 2025), this may help explain the calorie labelling regulations’ limited impact on population behaviour, specifically with respect to calories purchased and consumed OOH (Kalbus et al., 2025; Polden et al., 2024b).

The share of restaurants displaying calories in this study is well below the 17% of outlets that were estimated to be affected by the policy (Department of Health and Social Care, 2020). This may be explained by compositional differences between the physical and digital food environment. However, a recent analysis found an even higher share of large restaurant chains, which are required to display calories, on Just Eat in the UK (22%) (Huang et al., 2025b). Given the high overlap between restaurants listed on Just Eat, Deliveroo and Uber Eats (Kalbus et al., 2023), the share of these chains is likely to be similar for these services. Instead, compliance with the regulations may explain the lower-than-expected provision of calorie labelling. Compliance with the English calorie labelling regulations is incomplete, with most businesses only partially following relevant guidance (Polden et al., 2024a), and enforcement mechanisms are lacking (Essman et al., 2025). While no research to date evaluated compliance in the digital OOH food sector in England, international research indicates that compliance online may also be imperfect (Vanderlee et al., 2023a). For instance, Greenthal et al. (2023) reported that on third-party delivery platforms in the US, 12–58% of restaurant chains required to display calorie labels did so, which was considerably lower than on restaurant websites (95%). An investigation of online menus from restaurant chains required to display calories in Australia also found lower compliance among delivery platforms than on restaurants’ own websites (Cassano et al., 2024).

In addition to the low availability of calorie labelling overall, our findings highlight substantial variation by area deprivation. Specifically, residents in more deprived areas are facing a double burden of a high and increasing exposure to restaurants selling predominantly unhealthy food overall (Huang et al., 2024), while relatively fewer of these provide calorie labelling. This variation likely reflects the differential food environment composition by area deprivation of large chains, which are required to display calories, and small and independent restaurants, which are not subject to the calorie labelling regulations (Bite Back, 2024). In turn, information provision and the limited benefits of the policy observed here and elsewhere are concentrated in less deprived areas. Thus, the current policy’s focus on large businesses only while exempting smaller businesses, which disproportionally locate in more deprived areas, is a design limitation which may widen dietary inequalities.

Our observation of the presence of calorie labelling in Scotland and Wales, where labelling remains voluntary, corresponds to earlier observations of spillover effects. Vanderlee et al. (Vanderlee et al., 2023b) investigated the availability of calorie labels among restaurant chains in Canadian jurisdictions with and without mandatory calorie labelling on an online food delivery platform. They found that although calorie labelling was more prevalent in provinces where labelling was mandatory, chains also provided labelled menus in provinces where labelling was voluntary, albeit to a smaller degree (Vanderlee et al., 2023b). In our study, we observed that the provision of labelling was comparable in regions where it was mandatory (England) and where it was not (Scotland & Wales), suggesting that cross-border chains treated the calorie labelling regulations as a national policy.

The observed overall decrease in calories on menus, albeit small, may suggest positive, structural change which does not rely on consumer behaviour change (Garrott et al., 2024). This reduction was not patterned by area deprivation, suggesting an equitable change in the food environment. However, this applies only to restaurants displaying calories, with residents in more deprived areas being exposed to relatively fewer of these restaurants, and less healthy food available overall (Huang et al., 2024). The difference in change in calorie content by restaurant type likely corresponds to the types of food and associated calorie content sold. For instance, a greater calorie reduction was observed for ‘Burger’ (-23 kcal/food item) than for ‘Healthy’ (-8 kcal/food item) restaurants, but the median calorie content of the latter was well below the former (600 vs 407 kcal in October 2023). However, findings by restaurant type need to be interpreted with caution due to restaurants tagging themselves.

### Limitations

Several limitations apply to this study. First, as no pre-policy data were available, this study examined changes over time from policy implementation rather than compared menus in a pre-post design. Second, Just Eat, the largest delivery platform in the UK (SLERP, 2024) was not included in this study due to technical difficulties implementing automated data collection tools. However, bias from missing restaurants may be limited as previous research identified substantial overlap in restaurants signed up to Just Eat, Deliveroo and Uber Eats in the UK (Kalbus et al., 2023). Third, we set a threshold of 10% labelled menu items grounded in data exploration to not determine restaurants as displaying calories that provide calorie information for very few packaged items rather than their core menu. This definition may not be accurate enough in capturing large food businesses required to display calories. Fourth, as no information on an item’s serving size was available, we were not able to assess changes in calorie content adjusted for volume and can therefore not rule out if calorie reductions resulted from reducing portion sizes. Finally, our analysis considers the supply side only, and is not weighted by restaurant sales. In turn, while a comprehensive reflection of the supply on two major food delivery services, our analysis may not correspond to consumers’ exposure to calorie labelling and purchasing when using online food delivery.

### Implications for research and policy

The calorie labelling policy’s current focus on large restaurants may exacerbate dietary inequalities as people living in more deprived areas may be less exposed to nutritional information on OOH food. Stakeholders may therefore consider expanding mandatory calorie labelling to small- and medium-sized businesses, which may also encourage these businesses to reduce calories on their menus. A previous modelling study suggests large additional benefits from widening the regulations’ scope beyond large businesses (Colombet et al., 2024).

The digital OOH food sector is an increasingly important means of accessing food for many in the UK (Moore et al., 2025). At the same time, it poses a significant public health concern because it currently predominantly facilitates access to unhealthy food (Huang et al., 2024). With the use of online meal delivery patterned by socio-economic status and associated with weight status (Cummins et al., 2024; Nago et al., 2014), regulatory frameworks are required to improve nutritional quality in the sector to promote health and prevent the widening of dietary health inequalities. Approaches to monitor nutritional quality in the digital OOH food sector are emerging (Huang et al., 2024). Future research should build on these to understand how the sector is affected by policies such as the calorie labelling regulations and to identify priority areas for interventions.

Finally, while the overall reduction in calories may indicate a positive, structural change, further research is needed to understand how potential benefits of calorie reduction are distributed across the population.

### Conclusions

This study, using a complete online menu dataset from two major food delivery services in Great Britain, found that the share of restaurants displaying calories in Great Britain was low and falling. Overall, poorer neighbourhoods were doubly disadvantaged – not only did poorer areas have greater exposure to restaurants available for online delivery, but these areas also had the lowest proportion of restaurants displaying calorie labels. This suggests that the calorie labelling regulations may be poorly targeted, potentially widening inequalities in dietary health. However, observed reductions in calorie content over time may indicate positive structural change which does not rely on consumer behaviour change. Policy makers and researchers should consider how inequalities in the policy’s focus may translate to inequalities in dietary health.

## Data Availability

Due to its nature, we cannot share the data used in this study. However, the automated tools used for data collection are publicly available and referenced.

## Notes

### Competing Interest Statement

The authors have declared no competing interest.

### Funding Statement

This study is funded by the National Institute for Health and Care Research (NIHR) School for Public Health Research (SPHR) (Grant Reference Number NIHR 204000). JA is supported by the Medical Research Council (grant numbers MC_UU_00006/7). The views expressed are those of the authors and not necessarily those of the NIHR or the Department of Health and Social Care.

