## Appendix A for "How did labelling provision on menus for online food delivery change following England’s calorie labelling regulations?"

### Appendix A: Description of deduplication of restaurants

#### De-duplication of Restaurants

Some restaurants are listed on both delivery services (and deliver to the same neighbourhoods). We identified duplicated outlets to avoid double-counting them in the analysis.

Deduplication (record linkage) was performed for each neighbourhood (Lower layer Super Output Areas for England/Wales, Data Zones for Scotland) separately, i.e. all outlets from one delivery platform were linked to those from the other within each LSOA they deliver to. Using coordinates provided with the datasets, we calculated the distance (in m) between each record pair (using R package sf [1]). We allowed a threshold of 300 m for true duplicates to be apart, as coordinates may be recorded differently on both platforms (data exploration showed that true duplicates are no further apart in space as 300 m). We then processed the outlets' names to contain only letters and numbers. As many outlet names contained their location at the end of their name (e.g., 'pizzaria Camden', we extracted the first word of the name excluding common words such as 'restaurant', 'chicken' or 'the'. We calculated string similarity (using R package stringdist [2], method 'osa') for the first word excluding common words and the full string. We then classified record pairs duplicates when at least one of the following criteria was met:

- > 80% full-name similarity & 100% first-word similarity
- At least 90% full-name similarity & at least 30% first-word similarity
- Distance < 150 m and 100% first-word similarity
- Distance < 100 m & 100% first-word similarity & at least 20% full-name similarity
- Distance < 100 m & at least 60% full-name and first-word similarity, respectively
- Distance < 50 m & at least 80% full-name similarity

To assess the performance of this deduplication code, we determined duplicates as described above for outlet record pairs for randomly drawn ten neighbourhoods in England (n=7), Scotland (n=2) and Wales (n=1) which were less than 300 m apart in space from the June 2023 data. This sample consisted of 9,892 record pairs which comprised 1,276 and 820 unique outlets on Deliveroo and Uber Eats, respectively. The average sensitivity (= percentage of correctly identified duplicates out of true duplicates) was 94.78%, and the average specificity (= percentage of correctly identified non-duplicate pairs out of true non-duplicates) was 99.44%. The average positive predictive value (= percentage of correctly identified duplicates out of those identified as duplicates) was 95.78%, and the negative predictive value (= percentage of correctly identified non-duplicates out of those identified as non-duplicates) was 98.69%. Average accuracy (= percentage of correctly classified out of all pairs) was 98.29%.

Because one platform contains more restaurants than the other and its restaurants deliver to more areas than through the other platform (median 123 vs 67 neighbourhoods), we further took care to reduce errors in the record linkage (deduplication). Hence, we removed restaurants from the second platform with fewer

total outlets: e.g., 47,074 outlets flagged as duplicates on this platform compared to 49,675 outlets on the other – of which more are likely to be incorrect.
We identified duplicated records for each neighbourhood separately. Therefore, for the area-level analysis (analysis 1), we removed duplicates specific to this (delivery) area. By doing this, we account for restaurants having different delivery radii through the two platforms, and only removed duplicates where an area had access to the same restaurant through both platforms.
