## Appendix B for "How did labelling provision on menus for online food delivery change following England’s calorie labelling regulations?"

#### Analysis 1

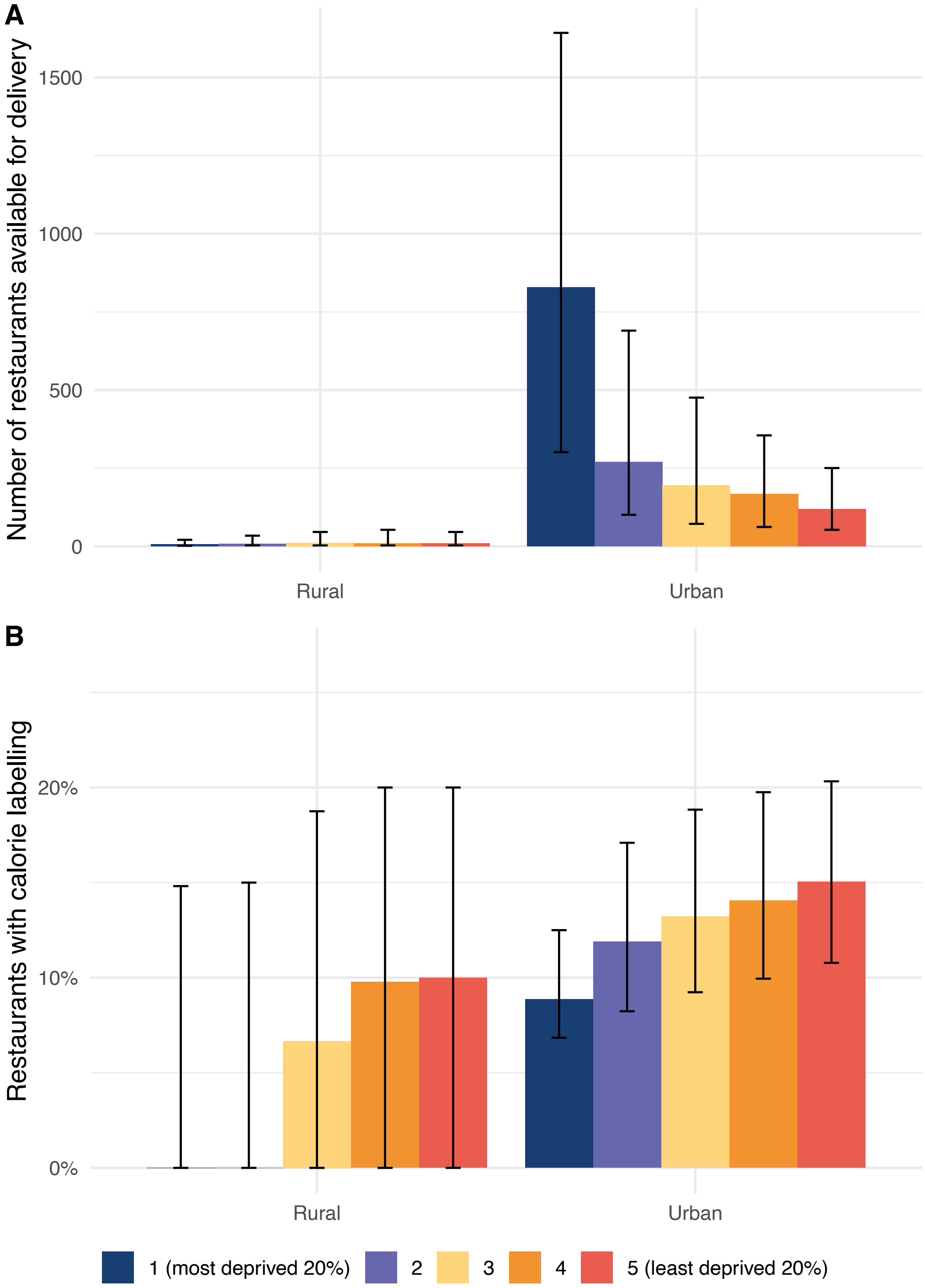


**Figure B.1. Number of all restaurants for delivery and % restaurants displaying calories by area deprivation quintile and urban status in October 2023.** Area deprivation was approximated through the Townsend Index. Medians per neighbourhood (LSOA in England and Wales/Data Zone in Scotland) are displayed. Error bars indicate the interquartile range.

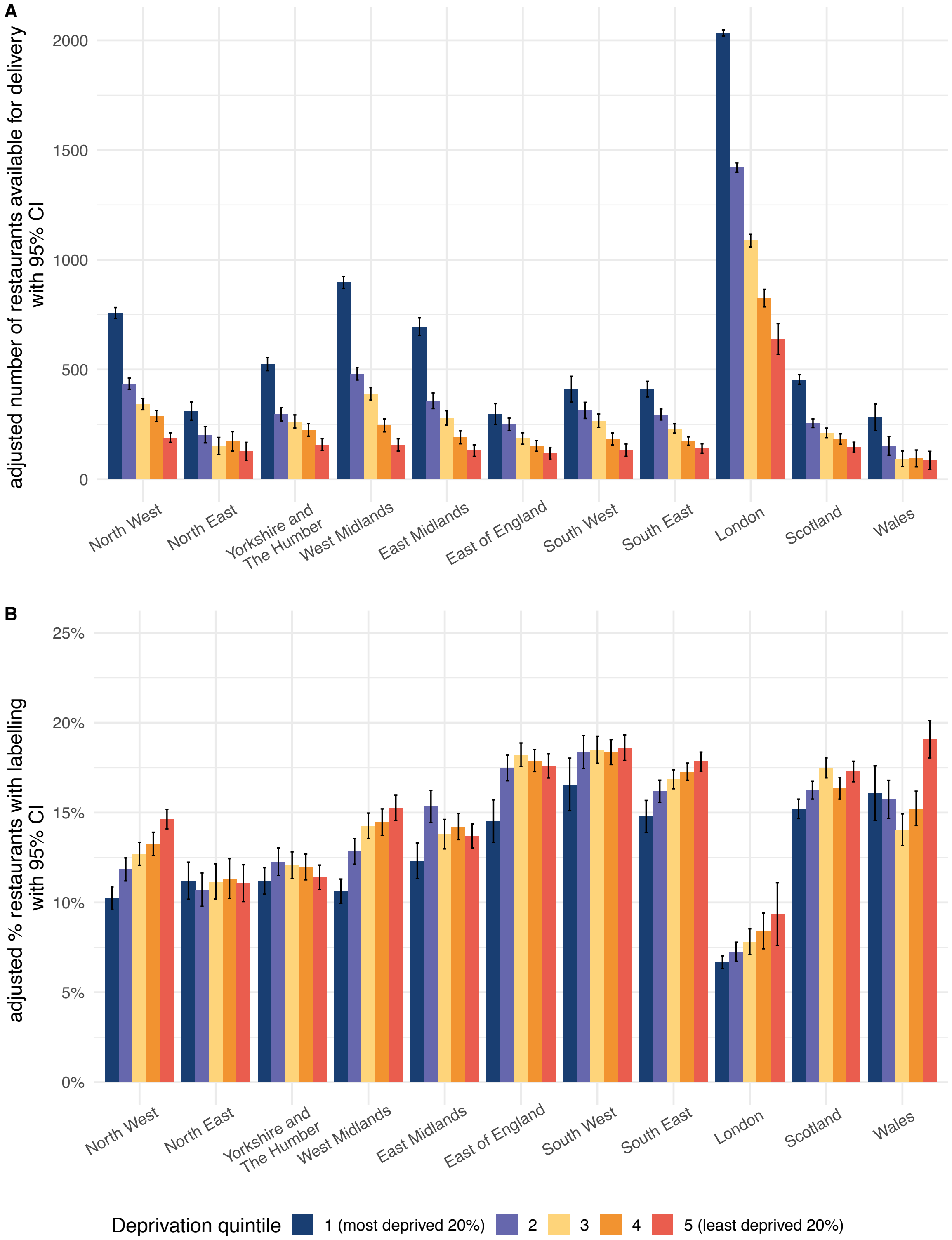


**Figure B.2. Adjusted number of restaurants available for delivery overall (A) and % restaurants displaying calories (B) by area deprivation and region in October 2023.** Area deprivation was approximated through the Townsend Index. Average predictions were derived from linear regression models adjusted for urban status, population density, and an interaction between region and deprivation quintile. Error bars indicate 95% confidence intervals.

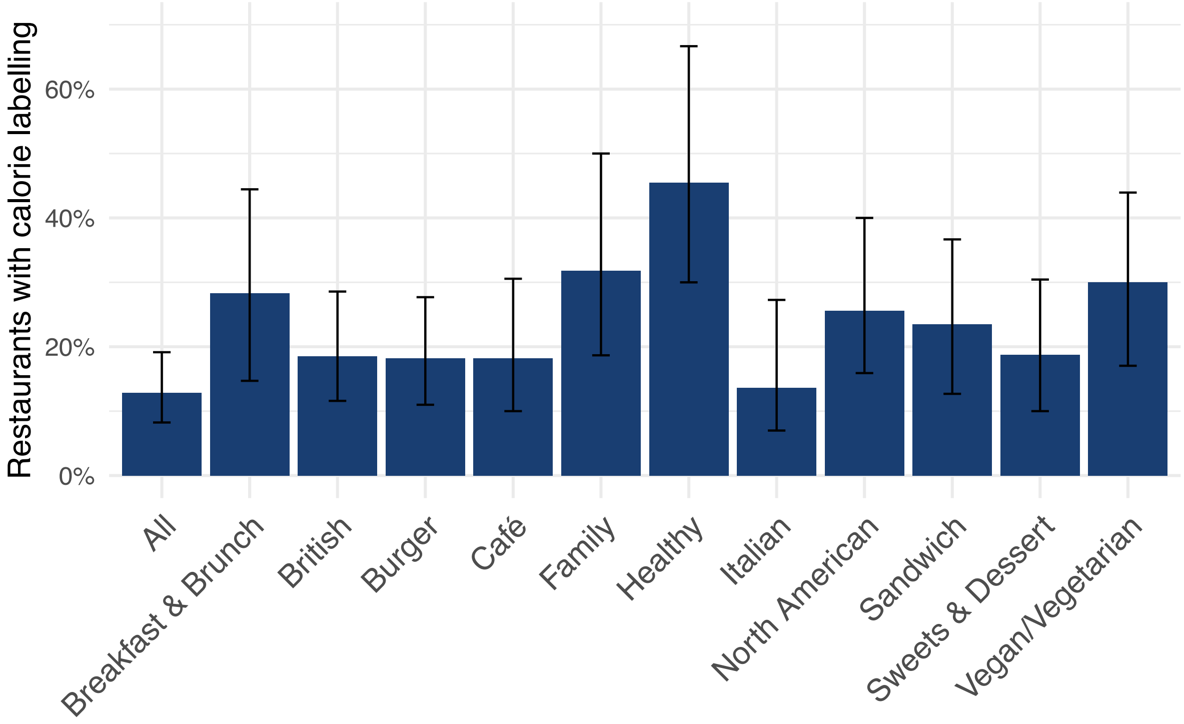


**Figure B.3. Calorie labelling provision by restaurant type in June 2023.** Type was determined through restaurant tags on the delivery websites. % restaurants with calorie labelling denotes the median percentage of restaurants delivering to a neighbourhood (LSOA in England and Wales/Data Zone in Scotland) with calorie labelling. Error bars indicate the interquartile range.

**Table B.1**. Calorie labelling provision in neighbourhoods

| | | June 2022 | October 2022 | February 2023 | June 2023 | October 2023 |
|---|---|---|---|---|---|---|
| Median number of available restaurants & % displaying calories (IQR) | # | 158 (51 to 425) | 172 (54 to 462) | 177 (56 to 477) | 187 (59 to 499) | 199 (60 to 551) |
| | % | 14.0 (8.8 to 20.9) | 14.2 (9.1 to 20.7) | 13.5 (8.6 to 20.0) | 12.8 (8.2 to 19.1) | 12.1 (7.9 to 18.0) |
| Median number of available restaurants & % displaying calories by deprivation quintile (IQR) | | | | | | |
| 1 – most deprived 20% | # | 628 (223 to 1,180) | 685 (243 to 1,298) | 696 (251 to 1,360) | 733 (262 to 1,438) | 820 (294 to 1,633) |
| | % | 10.2 (7.3 to 14.3) | 10.3 (7.4 to 14.5) | 9.8 (7.1 to 13.9) | 9.1 (6.8 to 13.1) | 8.9 (6.8 to 12.5) |
| 2 | # | 202 (78 to 501) | 218 (83 to 545) | 225 (86 to 561) | 236 (89 to 594) | 257 (92 to 654) |
| | % | 13.8 (8.9 to 20.0) | 13.9 (9.3 to 19.9) | 13.3 (8.9 to 19.1) | 12.4 (8.4 to 18.4) | 11.8 (8.1 to 17.0) |
| 3 | # | 134 (47 to 349) | 144 (50 to 381) | 149 (51 to 391) | 158 (53 to 407) | 167 (54 to 440) |
| | % | 15.2 (10.1 to 22.1) | 15.2 (10.3 to 21.8) | 14.5 (9.7 to 20.9) | 13.9 (9.2 to 20.0) | 13.0 (8.8 to 18.8) |
| 4 | # | 95 (29 to 229) | 103 (30 to 252) | 105 (31 to 258) | 111 (34 to 273) | 117 (33 to 298) |
| | % | 15.9 (10.4 to 23.4) | 15.9 (10.5 to 23.1) | 15.0 (9.8 to 21.9) | 14.6 9.4 to 21.1) | 13.7 (9.0 to 19.8) |
| 5 – least deprived 20% | # | 71 (20 to 161) | 77 (21 to 174) | 79 (21 to 179) | 83 (22 to 190) | 84 (21 to 204) |
| | % | 16.7 (10.1 to 23.6) | 16.7 (10.5 to 23.5) | 15.8 (9.9 to 22.2) | 15.5 (9.7 to 21.5) | 14.4 (9.3 to 20.2) |
| Neighbourhoods with online food delivery (%) | | 36,884 (88.4%) | 36,973 (88.6%) | 37,140 (89.0%) | 36,999 (88.7%) | 37,094 (88.9%) |
| Total number (unique) restaurants | | 98,289 | 103,184 | 106,836 | 110,736 | 114,256 |
| Number of (unique) restaurants displaying calories (%) | | 17,746 (18.1%) | 18,204 (17.6%) | 18,258 (17.1%) | 18,640 (16.8%) | 18,823 (16.5%) |

Neighbourhood = Lower layer Super Output Area (England & Wales) and Data Zone (Scotland) boundaries from 2011; area deprivation approximated through Townsend computed from Census 2021 (England & Wales) and 2022 (Scotland)

**Table B.2**. Provision of calorie labelling by region

| Region | Median number of available restaurants (IQR) | | Median % of restaurants showing calorie labels | |
|---|---|---|---|---|
| | June 2022 | October 2023 | June 2022 | October 2023 |
| All of England | 177 (68 to 465) | 223 (80 to 619) | 14.0 (8.8 to 20.9) | 12.1 (7.9 to 18.0) |
| North West | 150 (69 to 359) | 194 (83 to 517) | 12.7 (8.9 to 18.1) | 10.2 (7.8 to 15.1) |
| North East | 110 (43 to 181) | 125 (47 to 220) | 12.1 (7.1 to 14.9) | 10.8 (8.1 to 13.6) |
| Yorkshire and The Humber | 146 (67 to 294) | 184 (71 to 365) | 12.0 (8.6 to 18.7) | 10.0 (7.7 to 15.5) |
| West Midlands | 208 (81 to 449) | 277 (105 to 6140 | 12.6 (10.3 to 19.4) | 11.3 (9.3 to 15.9) |
| East Midlands | 98 (29 to 317) | 112 (34 to 392) | 15.1 (10.3 to 21.0) | 13.3 (8.5 to 17.6) |
| East of England | 141 (39 to 223) | 170 (43 to 260) | 20.3 (14.0 to 25.9) | 16.3 (11.4 to 21.5) |
| South West | 93 (24 to 228) | 113 (33 to 301) | 20.0 (15.0 to 25.5) | 17.6 (12.1 to 20.7) |
| South East | 139 (57 to 235) | 170 (70 to 297) | 18.8 (13.8 to 25.0) | 16.7 (12.1 to 20.7) |
| London | 1,011 (677 to 1,623) | 1,427 (916 to 2,312) | 7.3 (6.3 to 8.4) | 6.9 (6.1 to 7.7) |
| Scotland | 72 (23 to 299) | 87 (28 to 401) | 15.4 (11.1 to 22.6) | 14.2 (10.5 to 20.0) |
| Wales | 36 (15 to 117) | 41 (17 to 152) | 15.5 to (10.4 to 25.6) | 13.7 (7.5 to 21.4) |

**Table B.3**. Provision of calorie labelling by restaurant type

| Restaurant type | Median number of available restaurants (IQR) | | Median % of restaurants displaying calorie labels | |
|---|---|---|---|---|
| | June 2022 | June 2023 | June 2022 | June 2023 |
| North American | 33 (12 to 95) | 40 (14 to 116) | 29.4 (18.3 to 44.4) | 25.6 (15.9 to 40.0) |
| Breakfast & brunch | 18 (7 to 44) | 23 (9 to 56) | 15.5 (29.2 to 50.0) | 28.3 (14.7 to 44.4) |
| British | 26 (10 to 68) | 32 (12 to 83) | 16.7 (9.9 to 27.3) | 18.5 (11.6 to 28.6) |
| Burger | 36 (12 to 106) | 43 (14 to 129) | 20.5 (12.5 to 32.6) | 18.2 (11.0 to 27.7) |
| Café | 21 (8 to 56) | 28 (10 to 75) | 19.2 (11.1 to 33.3) | 18.2 (10.0 to 30.6) |
| Family | 15 (6 to 41) | 20 (7 to 58) | 33.3 (20.0 to 52.2) | 31.8 (18.7 to 50.0) |
| Healthy | 8 (3 to 25) | 11 (4 to 31) | 50.0 (33.3 to 83.3) | 45.5 (30.0 to 66.7) |
| Italian | 25 (9 to 69) | 27 (10 to 78) | 15.4 (7.4 to 30.0) | 13.6 (7.0 to 27.3) |
| Sandwich | 21 (8 to 62) | 28 (10 to 83) | 25.6 (14.2 to 42.9) | 23.5 (12.7 to 36.7) |
| Sweets & desserts | 28 (10 to 80) | 36 (12 to 103) | 20.0 (10.6 to 33.3) | 18.8 (10.0 to 30.4) |
| Vegan / Vegetarian | 38 (12 to 111) | 47 (14 to 136) | 30.2 (18.2 to 44.7) | 30.0 (17.0 to 43.9) |

### Analysis 2

**Table B.4**. Provision of calorie labelling among restaurants displaying calories, median (IQR)

| | June 2022 | October 2022 | February 2023 | June 2023 | October 2023 |
|---|---|---|---|---|---|
| All items | | | | | |
| n | 62 (36 to 90) | 63 (38 to 91) | 65 (42 to 89) | 65 (40 to 90) | 67 (40 to 95) |
| n with calorie information | 46 (24 to 74) | 49 (25 to 77) | 48 (26 to 74) | 45 (24 to 76) | 44 (23 to 80) |
| % with calorie information | 79.4 (63.4 to 96.7) | 79.7 (63.0 to 95.6) | 79.7 (63.2 to 95.8) | 77.9 (60.7 to 95.3) | 76.3 (56.1 to 93.2) |
| Food items | | | | | |
| n | 47 (26 to 69) | 47 (27 to 70) | 46 (28 to 65) | 44 (27 to 68) | 47 (27 to 70) |
| n with calorie information | 37 (19 to 64) | 40 (20 to 66) | 39 (20 to 60) | 37 (20 to 61) | 36 (18 to 61) |
| % with calorie information | 93.1 (78.9 to 99.3) | 93.3 (76.9 to 99.2) | 91.6 (74.3 to 99.3) | 90.9 (72.6 to 99.3) | 87.5 (68.6 to 98.1) |
| Drink items | | | | | |
| n | 13 (8 to 23) | 14 (8 to 22) | 15 (8 to 22) | 14 (8 to 23) | 14 (8 to 24) |
| n with calorie information | 7 (2 to 13) | 7 (1 to 12) | 8 (2 to 14) | 8 (2 to 13) | 7 (2 to 13) |
| % with calorie information | 59.8 (23.2 to 98.0) | 55.3 (22.2 to 98.2) | 64.2 (22.3 to 98.7) | 63.6 (20.8 to 98.6) | 63.6 (22.0 to 98.7) |

**Table B.5**. Provision of calorie labelling among restaurants displaying calories by area deprivation quintile, weighted median % labelled menu items (IQR) in October 2023

| Area deprivation quintile | All items | Food items | Drink items |
|---|---|---|---|
| 1 – most deprived 20% | 77.1 (58.5 to 93.6) | 89.8 (70.8 to 98.6) | 60.0 (17.9 to 100.0) |
| 2 | 77.8 (61.4 to 94.2) | 90.6 (71.9 to 98.7) | 62.5 (22.2 to 100.0) |
| 3 | 78.0 (62.9 to 94.7) | 90.9 (73.3 to 98.6) | 61.9 (25.0 to 100.0) |
| 4 | 78.1 (63.3 to 94.9) | 91.0 (74.1 to 98.6) | 60.9 (25.0 to 100.0) |
| 5 – least deprived 20% | 78.1 (63.7 to 94.9) | 91.3 (75.0 to 98.6) | 60.0 (25.0 to 100.0) |

IQR = interquartile range. Area deprivation was approximated using the Townsend Deprivation Index and considering each neighbourhood-restaurant combination (LSOAs in England and Wales, Data Zones in Scotland).

**Table B.6**. Provision of calorie labelling among restaurants displaying calories by restaurant type

| Restaurant type | # food outlets (June 2023) | Median % of labelled food items (IQR) | | Median % of labelled drink items (IQR) | |
|---|---|---|---|---|---|
| | | June 2022 | June 2023 | June 2022 | June 2023 |
| North American | 7,927 | 96.0 (82.2 to 99.4) | 93.6 (81.0 to 99.5) | 79.6 (22.9 to 98.5) | 76.4 (16.6 to 98.5) |
| Breakfast & brunch | 5,847 | 93.3 (86.3 to 99.4) | 92.9 (81.5 to 99.2) | 58.2 (24.8 to 95.8) | 65.9 (25.9 to 94.1) |
| British | 3,867 | 98.9 (84.5 to 99.4) | 97.5 (85.7 to 99.4) | 48.7 (30.0 to 89.7) | 74.4 (26.8 to 98.4) |
| Burger | 5,740 | 99.3 (92.3 to 99.6) | 98.6 (87.9 to 99.4) | 77.4 (34.5 to 98.1) | 76.8 (35.4 to 98.3) |
| Café | 4,170 | 90.0 (81.7 to 99.0) | 86.0 (74.0 to 97.5) | 35.8 (17.2 to 87.5) | 39.9 (19.2 to 80.6) |
| Family | 3,344 | 88.1 (75.7 to 98.9) | 85.5 (68.4 to 95.5) | 49.7 (11.4 to 84.4) | 48.9 (21.5 to 90.3) |
| Healthy | 4,092 | 97.5 (89.7 to 98.8) | 94.4 (81.2 to 98.6) | 88.5 (36.2 to 97.1) | 76.0 (31.9 to 98.7) |
| Italian | 2,941 | 85.8 (75.8 to 93.4) | 84.6 (64.7 to 92.3) | 32.3 (0 to 66.1) | 39.0 (15.3 to 95.4) |
| Sandwich | 6,170 | 96.0 (86.6 to 98.2) | 94.1 (82.9 to 98.6) | 90.8 (29.9 to 98.3) | 76.5 (35.1 to 98.8) |

| | | | | | |
|---|---|---|---|---|---|
| Sweets & desserts | 4,707 | 87.8 (81.6 to 94.8) | 85.7 (70.5 to 95.1) | 58.7 (0 to 97.7) | 56.2 (16.4 to 97.6) |
| Vegan / Vegetarian | 10,392 | 93.7 (82.0 to 99.2) | 93.5 (76.4 to 99.4) | 66.4 (31.9 to 98.3) | 73.1 (21.3 to 98.5) |

IQR = interquartile range. Estimates are weighted by the number of neighbourhoods (LSOAs in England and Wales, Data Zones in Scotland) the restaurant delivers to. Note that tags are not mutually exclusive, so the same restaurant could appear in more than one type.

### Analysis 3

**Table B.7**. Weighted median calorie content (kcal, IQR) by whether an item was on the menu continuously or not among restaurants displaying calories

| | Food items | | Drink items | |
|---|---|---|---|---|
| | Continuously on the menu | New/removed | Continuously on the menu | New/removed |
| June 2022 | 474 (383 to 656) | 534 (377 to 714) | 63.7 (13.2 to 117) | 62.8 (30.0 to 110) |
| October 2023 | 459 (385 to 633) | 521 (373 to 694) | 62.5 (9.02 to 123) | 76.1 (29.7 to 120) |

**Table B.8**. Calorie content of menu items among restaurants displaying calories by area deprivation quintile, weighted median kcal (IQR)

| Area deprivation quintile | All items | | Food items | | Drink items | |
|---|---|---|---|---|---|---|
| | 2022 | 2023 | 2022 | 2023 | 2022 | 2023 |
| 1 – most deprived 20% | 437 (303 to 605) | 419 (282 to 559) | 505 (387 to 690) | 481 (375 to 629) | 63 (28 to 100) | 59 (14 to 97) |
| 2 | 439 (316 to 616) | 414 (293 to 579) | 505 (387 to 700) | 497 (373 to 649) | 63 (28 to 95) | 58 (15 to 95) |
| 3 | 440 (320 to 621) | 414 (298 to 582) | 505 (387 to 702) | 498 (372 to 667) | 63 (28 to 93) | 58 (17 to 93) |
| 4 | 439 (320 to 625) | 413 (300 to 585) | 505 (384 to 700) | 500 (371 to 675) | 63 (28 to 89) | 58 (18 to 92) |
| 5 – least deprived 20% | 435 (320 to 624) | 409 (300 to 586) | 504 (382 to 700) | 498 (367 to 676) | 63 (28 to 88) | 57 (20 to 90) |

June 2022 and October 2023. Area deprivation was approximated using the Townsend Deprivation Index and considering each neighbourhood-restaurant combination (LSOAs in England and Wales, Data Zones in Scotland).

**Table B.9**. Weighted median calorie (kcal) content (IQR) of menu items among restaurants displaying calories by restaurant type

| | # restaurants | Food items | | Drink items | |
|---|---|---|---|---|---|
| Restaurant type | June 2023 | June 2022 | June 2023 | June 2022 | June 2023 |
| North American | 7,927 | 560 (422 to 712) | 570 (444 to 689) | 62.7 (21.4 to 97.0) | 52.4 (9.63 to 111) |
| Breakfast & brunch | 5,847 | 370 (337 to 483) | 374 (350 to 484) | 82.9 (42.1 to 98.6) | 87.8 (37.6 to 138) |
| British | 3,867 | 558 (474 to 705) | 471 (404 to 641) | 63.3 (7.4 to 75.1) | 64.9 (4.6 to 114) |
| Burger | 5,740 | 623 (474 to 739) | 600 (450 to 682) | 62.2 (12.6 to 83.8) | 73.5 (13.6 to 116) |
| Café | 4,170 | 377 (331 to 539) | 361 (342 to 464) | 87.1 (42.7 to 112) | 89.5 (37.0 to 140) |
| Family | 3,344 | 618 (464 to 755) | 585 (478 to 713) | 58.9 (26.4 to 71.2) | 54.1 (4.7 to 113) |
| Healthy | 4,092 | 415 (316 to 489) | 407 (355 to 489) | 62.8 (42.2 to 119) | 79.3 (44.5 to 119) |
| Italian | 2,941 | 666 (490 to 875) | 595 (503 to 780) | 59.6 (11.9 to 73.4) | 34.1 (1.8 to 82.8) |
| Sandwich | 6,170 | 360 (317 to 436) | 371 (349 to 438) | 59.9 (36.6 to 94.2) | 69.5 (36.7 to 124) |
| Sweets & desserts | 4,707 | 558 (436 to 787) | 545 (363 to 705) | 71.1 (36.2 to 114) | 69.8 (36.1 to 132) |
| Vegan / Vegetarian | 10,392 | 504 (420 to 700) | 478 (373 to 619) | 59.9 (26.6 to 95.7) | 64.0 (13.6 to 118) |
